# Excess respiratory, circulatory, neoplasm and other mortality rates during the Covid-19 pandemic in the EU and their implications

**DOI:** 10.1101/2025.03.07.25322542

**Authors:** Gabrielle Kelly, Stefano Petti, Norman Noah

**Affiliations:** School of Mathematics and Statistics, University College Dublin, Ireland; Department of Public Health and Infectious Diseases, Sapienza University, Rome, Italy; Department of Infectious Disease Epidemiology, London School of Hygiene and Tropical Medicine, London, U.K

## Abstract

Recently data has become available on excess mortality rates due to specific causes during the Covid-19 pandemic. Using the Eurostat database from 2016 to 2021, deaths attributed to Covid-19 in 2020-2021 and all deaths, as well as deaths due to specific causes including diseases of the circulatory and respiratory systems, neoplasms and transport accidents, were tabulated for 33 European countries. From the Our World in Data database, vaccination rates, economic, health, demographic and government response stringency index variables were also tabulated. Among the key findings were using standardised rates: (1) With the exception of Central and Eastern European countries, in almost all countries, excess mortality rates due to diseases of the circulatory, respiratory systems and neoplasms were negative. Ireland had the lowest excess respiratory rates both years; (2) Significantly positive excess mortality rates appeared in Croatia, Cyprus, Malta and Turkey and were attributed to neoplasms, diabetes, dementia and ill-defined and unknown causes of mortality, respectively. Indications are that these were exacerbated by public health measures (apart from Turkey); (3) In regression analyses the human development index and vaccination rates were important explanations for all excess death rates; (4) Statistically significant positive or negative excess rates for a disease could be flags for increased rates in the future.

## Introduction

Much research has focused on the elderly or those who have preexisting health conditions as being at high risk of dying of Covid-19, with being male also a factor [1–2]. The prevalences of some health conditions, however, are often country specific. While it has been reported that a substantial increase in excess mortality largely coincided with a Covid-19 outbreak in each European country, the indicator did not specify the causes of death [3–4]. In addition to confirmed deaths, the indicator captured Covid-19 deaths that were not correctly diagnosed and reported, as well as deaths from other causes that might have been attributed to Covid-19. It also accounted for a possible reduction in deaths from road traffic accidents that did not occur due to restrictions on commuting or travel during the lockdown periods. In this paper we focus on characteristics that explain differences in Covid-19 and excess mortality rates between countries.

In a recently published paper [5], excess mortality rates in 2020 for a snapshot of 35 countries were examined. Residual mortality rates (RMR) (i.e. non-Covid-19 excess mortality) in 2020 were then calculated as excess mortality minus reported Covid-19 mortality rates. Differences in RMR are differences not attributed to reported Covid-19 and of the 35 countries about half the RMR’s were negative. Examination of excess mortality rates due to non-Covid-19 causes may provide insights into indirect outcomes of the pandemic other than Covid-19 mortality. Circulatory diseases accounted for close to one-third (32.4 %) of all deaths in the EU in 2021. The second most common cause was cancer (21.6%), followed by Covid-19 which accounted for 10.7% and then followed by respiratory diseases [6]. These causes of death are investigated here; the results may then be used to determine which preventive and medical-curative measures or which investment in research might be used in a future pandemic. Differences in excess mortality rates between countries are also informative as to economic, social and health system factors that may affect them as well as government policies during the pandemic.

## Methods

Data were obtained from the Eurostat database [7]. From this database, yearly data on causes of death for 32 countries, namely, all 27 EU Member States, EFTA countries (Iceland, Liechtenstein, Norway, Switzerland), the two candidate countries Serbia and Turkey, was available from 2016-2021 inclusive. Monthly data was unavailable for causes of death in 2021 and therefore attention is restricted here to yearly data. Eurostat statistics on the causes of death are based on the medical information provided in the death certificates. Causes of death are classified by the 86 causes in the European shortlist which is based on the International Classification of Diseases and Related Health Problems (ICD-10) codes in its tenth revision, corresponding to the immediate cause of death. Codes U071 (virus identified, deaths where Covid-19 has been confirmed by laboratory testing), U072 (virus not identified, denoted by CNID) and U_COV19_OTH (Covid-19 death not elsewhere defined) were used to identify whether a death was related to Covid-19 infection codes (introduced by the WHO when the outbreak of Covid-19 started). Covid-19 identified will be denoted by CID and otherwise by Covidother (U072+U_COV19_OTH).

Standardised death rates (SDR) are examined rather than crude, as the population structure strongly influences this indicator for broad age classes. Excess mortality rates are then calculated for all causes and for specific causes. In this article, the excess mortality rate 2020 and 2021 is defined and calculated (as by Eurostat), as the SDR from all causes minus the average annual SDR over the previous four years (2016-2019) before the pandemic.

We then examined excess death rates due to the causes: circulatory diseases (codes I00-I99), neoplasms (codes C00-D48), non-Covid-19 respiratory diseases (codes J00-J99), as well as transport accidents (codes V-Y85) for 32 countries in 2020 (Turkish data unavailable) and 33 in 2021. Assuming a Poisson distribution for counts, mortality rates were tested for statistical significance.

The excess non-Covid-19 mortality rate for each country was calculated as the excess death rate minus death rates due to Covid-19 (identified, non-identified, other) [5].

Another dataset was obtained from the OWID database [8]. Variables provided include the stringency index (SI) as defined below, population size, population density, median age, aged 65 or older, aged 70 or older, per-capita GDP (gdp), extreme poverty, female smokers, male smokers, hospital beds per thousand, life expectancy and human development index (HDI, a composite index measuring average achievement in three basic dimensions of human development from the United Nations Development Programme: life expectancy, education and gross national income per capita). More information is available in the codebooks. The SI proposed in [9], is an indicator of the measures taken by governments against Covid-19. This is a composite metric based on nine reaction indicators, such as school closures, workplace closures, and travel restrictions, rescaled to a number between 0 and 100 (100 being the most stringent). People vaccinated per hundred of the population was available from the database for each day 2020 and 2021. Excess mortality rates for specific causes were only available per year, so to allow for comparisons and correlations, mean vaccination rate for 2020 and 2021 were calculated for each country, as well as the maximum (max), minimum (min) and standard deviation (sd). The same procedure was carried out for the SI. Correlation and stepwise regression analyses were carried out relating excess mortality rates separately for each cause and for 2020 and 2021 to these explanatory variables. Spearman correlation coefficients were computed in all cases. Statistical significance denotes a p-value < 0.05. The percentage of variation explained by a regression model is denoted by R^2^.

The OECD’s definition of central and eastern European countries (CEECs) for the group of countries comprising Albania, Bulgaria, Croatia, Czechia, Hungary, Poland, Romania, Slovakia, Slovenia, Estonia, Latvia, and Lithuania was used [10].

## Results

Note that results concerning Liechtenstein will not be commented on due to its small population and subgroups sizes, where statistically significant results in rates may only reflect very small differences in absolute numbers. As the HDI was significantly positively correlated with vaccinations statistics in 2021, the latter often did not appear in the final regression model and relevant correlations are noted separately. Correlations with stringency statistics are also noted separately even if they are not statistically significant.

### Overall excess and non-Covid-19 excess mortality

Tables 1 and 2 show that overall excess mortality rates were negative for six countries in 2020 and for nine in 2021 but not significantly so in either year, while most countries were positive, many significantly so. Countries in eastern Europe had the highest excess mortality rates in 2021. In the stepwise regression analysis in 2020 the HDI (p=0.04) and age 70 or older (p=0.05) were negative and median age positively (p=0.01) associated with excess mortality (R^2^=0.48) and in 2021 female smokers (p=0.02) and the SI mean (p=0.08) were positively associated and HDI (p<0.0001), life expectancy (p<0.0001) and max vaccination rate (p=0.0001) were negatively associated (R^2^=0.96). When the SI mean was omitted from the final model, little changed (R^2^=0.95). The Figure displays the association between the excess mortality rate and the HDI. Seven of 32 countries had positive non-Covid-19 excess for 2020 while Ireland was the only country significantly negative. Eight countries had positive non-Covid-19 excess in 2021, and Bulgaria, Serbia and Turkey were significantly positive. Belgium, Ireland, Luxembourg and Sweden all had significantly negative non-Covid-19 excess in 2021. In the stepwise regression analysis in 2020 the HDI was negatively associated (p<0.0001) with non-Covid-19 excess (R^2^=0.52) and both mean vaccination rate (p=0.001) and HDI (p < 0.0001) were in 2021 (R^2^=0.76).

**Table 1.**
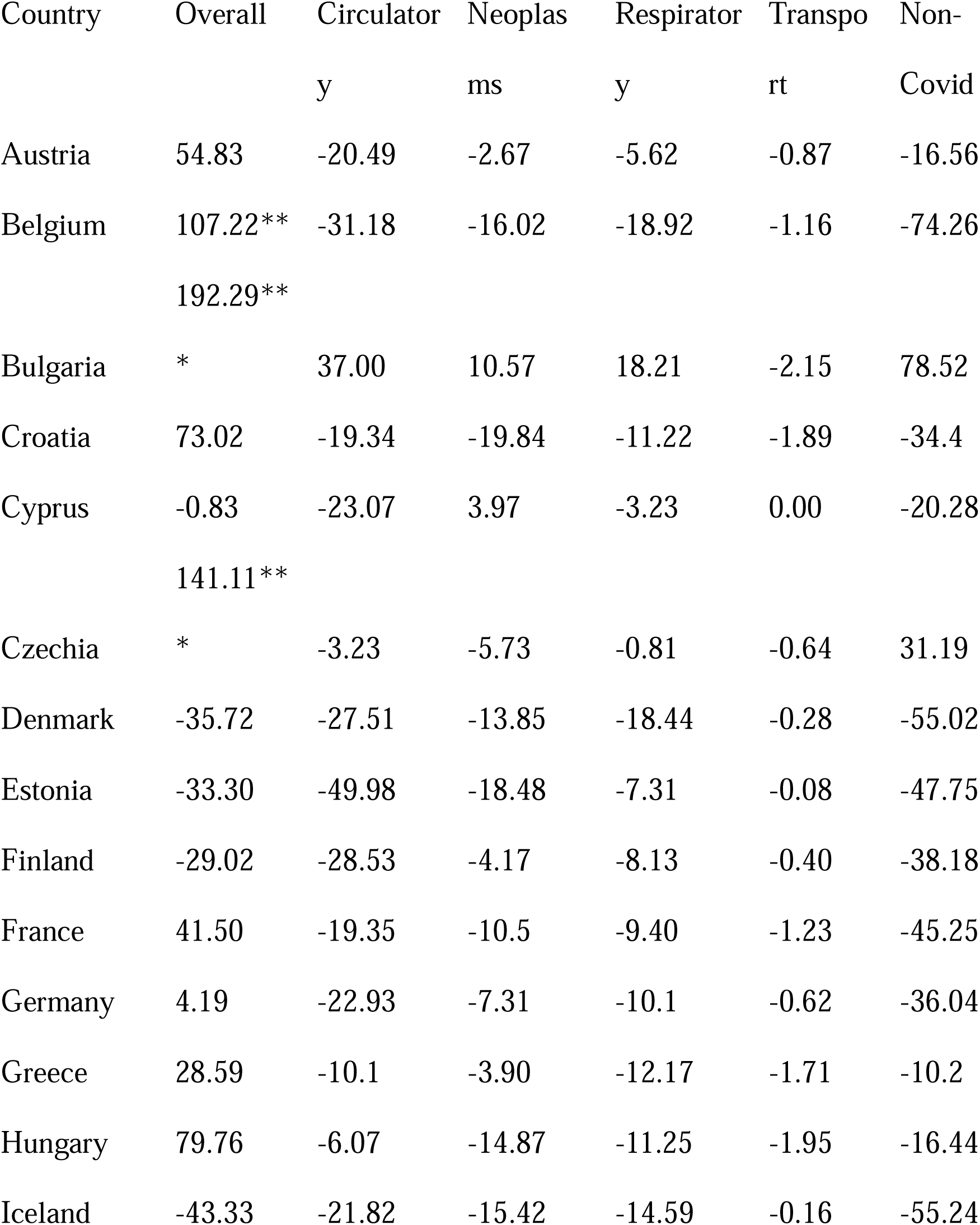
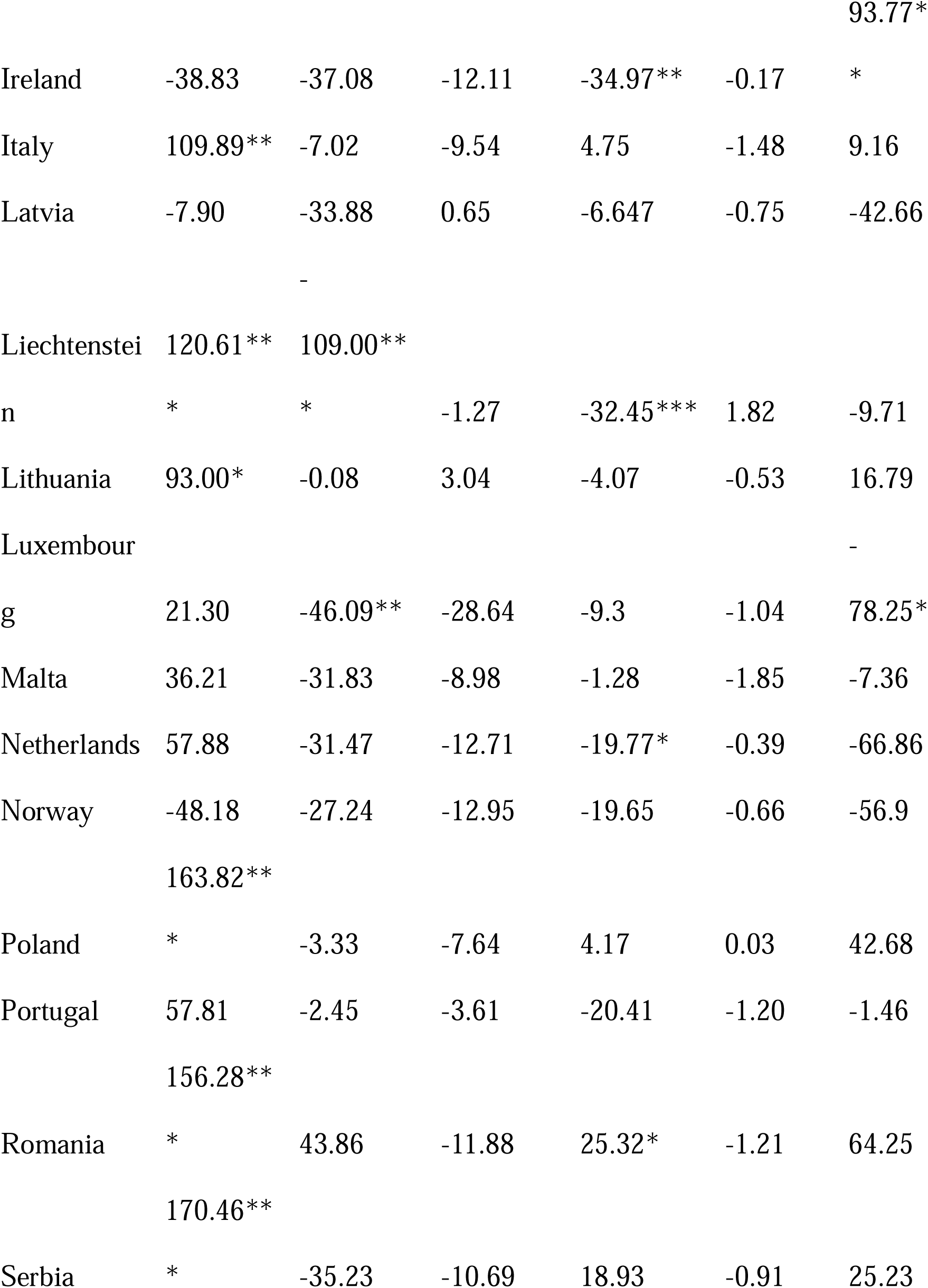
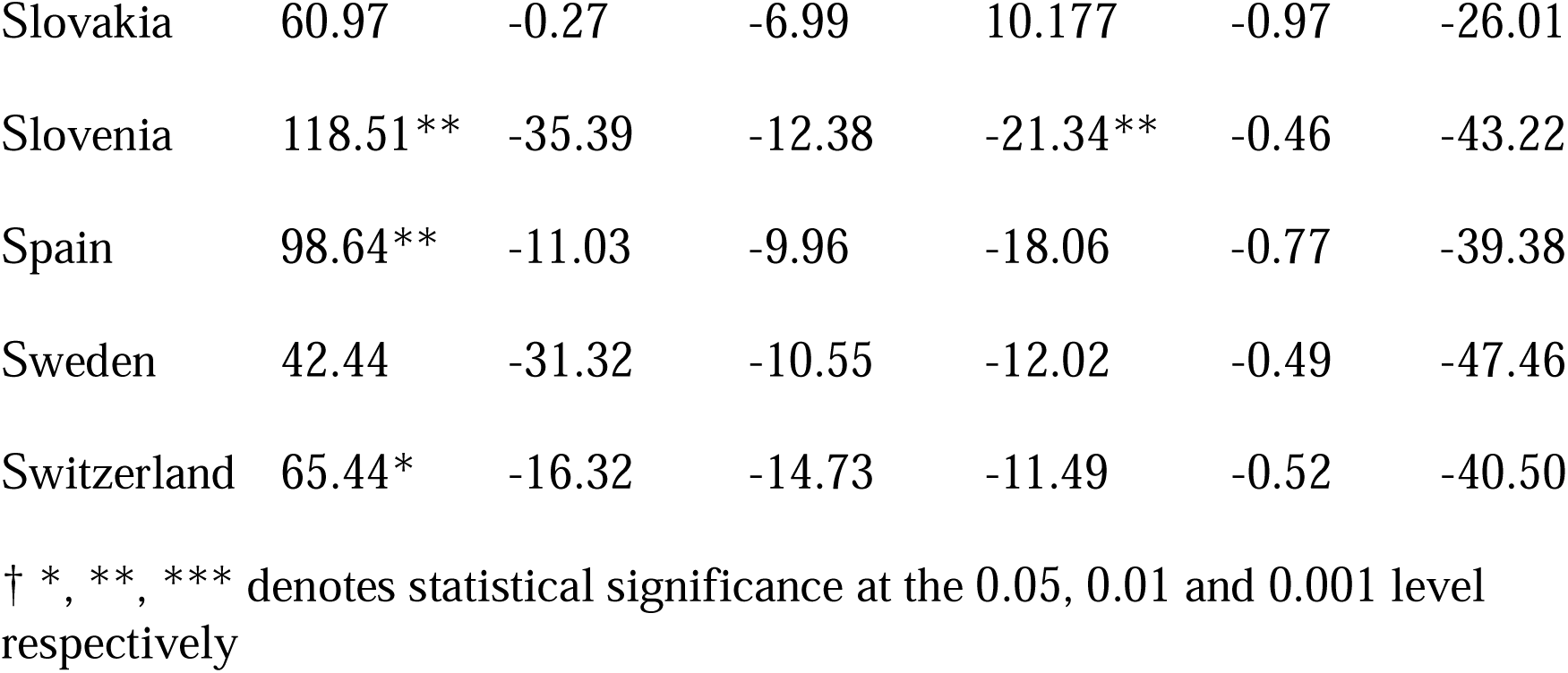
Excess standardised mortality rates per 100,000 population overall and for specific causes and non-Covid-19 excess mortality rates in 32 European countries in 2020 †.

**Table 2.** Excess standardised mortality rates per 100,000 population overall and for specific causes and non-Covid-19 excess mortality rates in 33 European countries in 2021 †.

| Country | Overall | Circulato<br>ry | Neoplas<br>ms | Respirato<br>ry | Transpo<br>rt | Non-Covid |
| --- | --- | --- | --- | --- | --- | --- |
| Austria | 42.97 | -38.20 | -8.83 | -15.40 | -0.76 | -42.58 |
| Belgium | -22.29 | -32.07 | -18.67 | -29.74** | -1.06 | -106.3** |
| Bulgaria | 537.45*<br>** | 128.29**<br>* | -5.40 | 29.82** | -2.15 | 163.91*<br>* |
| Croatia | 219.80*<br>** | -9.00 | -15.63 | -4.07 | -0.71 | 15.78 |
| Cyprus | 90.33* | -30.00 | 12.37 | -20.92 | -0.74 | 3.18 |
| Czechia | 230.80*<br>** | -32.77 | -16.02 | -7.49 | -1.10 | -22.73 |
| Denmark | -13.25 | -23.05 | -14.40 | -15.61 | -0.65 | -39.16 |
| Estonia | 146.88*<br>** | -10.79 | -18.02 | -0.35 | 0.28 | 20.47 |
| Finland | -16.39 | -31.35 | -6.18 | -8.77 | -0.25 | -31.78 |
| France | 26.30 | -17.11 | -14.75 | -12.50 | -0.87 | -50.98 |
| Germany | 29.03 | -26.15 | -12.18 | -14.97 | -0.77 | -44.09 |
| Greece | 125.51* | -5.72 | -6.17 | -8.71 | -1.38 | -1.69 |
|  | ** |  |  |  |  |  |
| Hungary | 218.66* | -14.63 | -26.60 | -15.94 | -1.13 | -39.53 |
|  | ** |  |  |  |  |  |
| Iceland | -63.68* | -31.84 | -21.46 | -16.64 | -1.18 | -65.38* |
| Ireland | -16.56 | -34.21 | -20.37 | -40.27*** | -0.53 | -101.8** |
| Italy | 55.73 | -21.27 | -15.53 | -10.46 | -0.51 | -26.79 |
| Latvia | 274.90* | 42.05 | -11.69 | -3.46 | 0.23 | 59.53 |
|  | ** |  |  |  |  |  |
| Liechtenste | -57.09 | - | -5.49 | -68.81*** | -1.57 | - |
| in |  | 65.59*** |  |  |  | 114.2*** |
| Lithuania | 234.16* | 7.13 | -14.66 | -9.00 | -1.75 | -3.18 |
|  | ** |  |  |  |  |  |
| Luxembour | -24.24 | - | -27.80 | -16.92 | -1.72 | -110.2** |
| g |  | 57.35*** |  |  |  |  |
| Malta | 14.01 | -46.08* | -22.96 | -1.58 | -3.18* | -29.88 |
| Netherland | 48.99 | -31.29 | -17.66 | -23.93** | -0.64 | -69.03 |
| s |  |  |  |  |  |  |
| Norway | -44.37 | -18.74 | -17.29 | -23.67* | -1.16 | -61.99 |
| Poland | 275.19* | 14.32 | -29.47 | 1.55 | -2.31 | 7.78 |
|  | ** |  |  |  |  |  |
| Portugal | 14.09 | -39.53* | -17.92 | -34.43*** | -1.04 | -88.72* |
| Romania | 395.52* | 131.12** | -29.04 | 40.12** | -0.96 | 172.78* |
|  | ** | * |  |  |  | ** |
| Serbia | 478.45* | 38.82 | -17.41 | 28.24* | -0.24 | 78.44 |
|  | ** |  |  |  |  |  |
| Slovakia | 371.90* | 33.86 | -31.95 | 73.44*** | -1.09 | 55.00 |
|  | ** |  |  |  |  |  |
| Slovenia | 56.99 | -51.30* | -25.99 | -28.41*** | 0.08 | -83.28* |
| Spain | 8.62 | -15.88 | -11.13 | -31.43*** | -0.53 | -65.82* |
| Sweden | -42.20 | -41.57* | -18.63 | -17.51* | -0.73 | -91.85** |
| Switzerland | -6.88 | -30.40 | -19.00 | -13.82 | -0.69 | -73.99* |
| d |  |  |  |  |  |  |
| Turkey | 269.33* | 37.12 | -22.68 | 48.78** | -2.50 | 112.35** |
|  | ** |  |  |  |  |  |

† \*, \*\*, \*\*\* denotes statistical significance at the 0.05, 0.01 and 0.001 level respectively.

**Figure:**
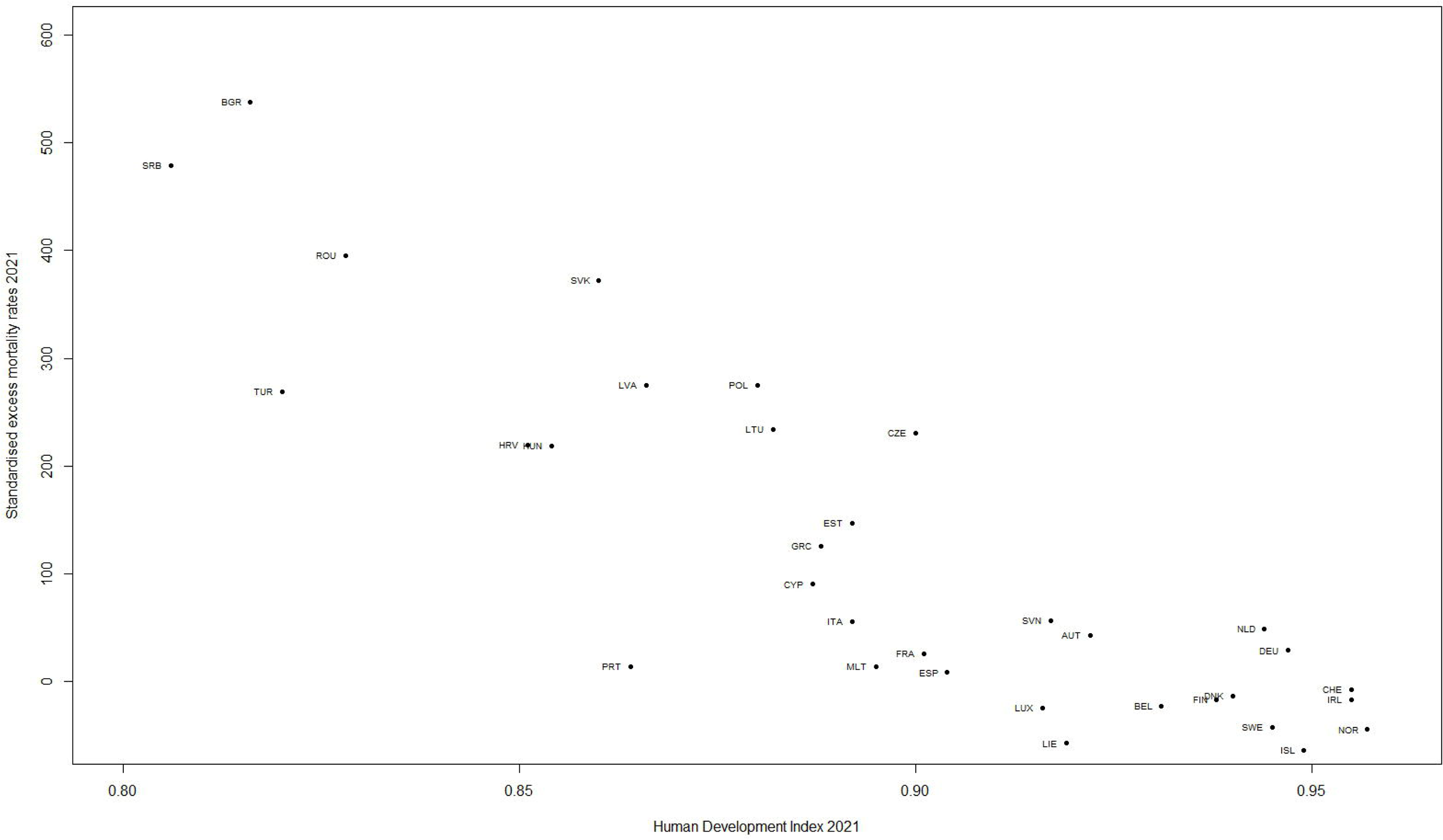
Standardised excess death rates per 100,000 of the population versus the human development index for 33 countries in 2021. Countries are labelled by their ISO code.

### Excess respiratory mortality

Only six countries had positive excess mortality due to respiratory diseases during 2020, these were all eastern European countries bar Italy, and Bulgaria, Romania and Serbia were again positive in 2021 - significantly so (Tables 1 and 2). Two countries had significantly negative excess respiratory mortality in 2020 - Ireland and Slovenia and ten countries had significantly negative excess respiratory mortality in 2021. In the stepwise regression analysis of excess respiratory mortality in 2020, HDI (p<0.001) was negatively, and hospital beds positively (p=0.05) related (R^2^=0.57). In 2021 HDI (p=0.003), max vaccination rate (p=0.005) and median age (p=0.02) were negatively associated while the sd vaccination rate was borderline negatively associated (p=0.06) and R^2^=0.70.

Note excess respiratory rates were negatively correlated with mean vaccination rate (0.61, p=0.0002), sd vaccination rate (0.56, p=0.0008) and max vaccination rate (0.54, p=0.0013). Excess respiratory rates were also negatively correlated with the mean SI in both 2020 and 2021 but not significantly.

### Excess circulatory mortality

Some CEEC’s had the highest mean circulatory diseases mortality rates 2016-2019. Luxembourg had significantly negative excess circulatory rates in 2020, while Bulgaria and Romania were the only countries positive. Seven CEEC countries were positive in 2021 with Bulgaria and Romania significantly so (Tables 1 and 2).

In the stepwise regression analysis in 2020, HDI was negatively (p<0.001) associated with excess circulatory (R^2^=0.45) and in 2021 the mean vaccination rate (p<0.001) and HDI (p=0.003) were negatively associated (R^2^=0.75). Note excess circulatory mortality was positively correlated with the mean, sd and max stringency index in 2020 and positively with the mean and negatively with sd and max in 2021 but not significantly.

### Excess neoplasms mortality

Some CEEC’s had the highest mean neoplasms mortality rates 2016-2019. Bulgaria, Cyprus, Latvia and Lithuania were positive for excess neoplasms in 2020, all other countries negative, while in 2021 all countries except Cyprus were negative. No country was significant in either year. In a stepwise regression in 2020, gdp (p=0.002) was negatively associated with excess neoplasms (R^2^=0.30) and in 2021 male smokers (p<0.001) and HDI (p=0.005) were both positively associated (R^2^=0.35). However, when Cyprus was omitted, male smokers was no longer significant in the model and age 70 and older (p<0.001) and female smokers (p<0.05) were positively associated, and median age negatively associated (p<0.01), with R^2^=0.52. Correlations between excess neoplasms in 2020 with the SI in 2020 and 2021 statistics were all very small and negative. Excess neoplasms in 2021 was negatively correlated with mean vaccination rate (0.54, p=0.001), sd vaccination rate (0.47, p=0.005) and max vaccination rate (0.47, p=0.006).

### Excess transport accidents mortality

Some CEEC’s had the highest mean transport accident mortality rates 2016-2019. Excess rates due to transport were almost all negative in 2020 (except Poland) and 2021 (except Estonia, Latvia and Slovenia) although none significantly so and rates were relatively low. In a stepwise regression of excess transport 2020, HDI (p<0.001) and sd SI (p=0.05) were positively associated, population density (p=0.04), female smokers (p=0.04) and life expectancy (p=0.04) negatively associated (R^2^=0.58). In 2021 age 70 older (p=0.0159), HDI (p=0.0194) was positively associated while population density (p=0.001) was negatively associated (R^2^=0.51). Population density was negatively associated with excess transport both years. This was driven by Malta with a population density almost three times any other country and with a low excess transport mortality rate. When Malta was omitted population density was not significant. Excess transport was negatively correlated with max SI (0.28, p=0.13), not significant in 2021 and all correlations with stringency statistics were negative in 2020 and 2021. Excess transport 2021 was not significantly correlated with vaccination statistics and all correlations were very small. Excess transport in 2021 was not significantly correlated with stringency 2020 statistics.

Results relating to CID and Covidother standardised mortality rates are in the Supplementary Material and have been discussed in part elsewhere [3,8].

## Discussion

Overall excess mortality in the EU has been extensively studied by for example in [3] and will only be briefly commented on here.

For most countries non-Covid-19 excess mortality rates in both 2020 and 2021 (Tables 1 and 2), were negative, meaning that the number of reported Covid-19 deaths is greater than the number of non-Covid-19 excess mortalities. This also indicates that non-Covid-19 deaths have been lower during the pandemic than expected, based on trends from previous years. Italy was one of the countries with positive non-Covid-19 excess in 2020 although not statistically significant. This may be due to the fact that the first deaths due to Covid-19 in Italy were on 22 February 2020 (before WHO guidelines, dated April 2020 to put Covid-19 as the underlying cause of death whenever Covid-19 was confirmed or even suspected were in place). A few more countries had positive non-Covid-19 excess in 2021, and some were significantly so. There are two factors that affect non-Covid-19 excess – the Covid-19 mortality rate as reported and the change in mortality rate due to other causes. Thus, non-Covid-19 excess may be positive or negative due to a combination of the following: (a) Covid-19 over-reporting (CovidNID and Other, large) -decreasing non-Covid-19 excess; (b) Covid-19 under-reporting of (CovidNID and Other, small) -increasing non-Covid-19 excess; (c) Increased mortality due to other causes -increasing non-Covid-19 excess; (d) Decreased mortality due to other causes -decreasing non-Covid-19 excess.

It is stated in [11], Covid-19 mortality can be estimated from excess deaths only when reliable mortality data are routinely available. This is difficult even then, however. Data on Covid-19 mortality and other reports show that only 1%-3% of all Covid-19 deaths were free of comorbidities [11–14]. Deaths attributed to Covid-19 cannot be attributed to other causes; consequently, rates of other causes of death may be reduced. This may have happened in Belgium that had a high rate of Covid-19 identified and other mortality (Table S1) and low non-Covid-19 excess (Tables 1,2). Belgium included deaths in care homes that were suspected, but not confirmed as Covid-19 cases. Belgium also has a high rate of care home occupancy relative to its population [5]. This supports the hypothesis that if Covid-19 is over-reported non-Covid-19 excess will be significantly negative.

Bulgaria and Romania stand out in 2020 and in 2021 as having significantly positive overall excess mortality with positive excess circulatory and respiratory diseases and Bulgaria also for neoplasms in 2020 -the only two countries positive for all three/or two of three in either 2020 or 2021. These results may indicate (b) or (c) above. Explanation (b) is not likely as both countries have comparatively high rates of Covid-19 and in the case of Romania, Covid-19 other is relatively low (Table S1). Romania and Bulgaria have the highest and second highest average (pre-Covid-19) rate of circulatory disease in the EU. It is reported they together with Croatia all entered the Covid-19 crisis with common problems, including workforce shortages and underdeveloped and underutilized preventive and primary care. The provision and uptake of non-Covid-19 health services was still affected negatively by the pandemic and was severely disrupted as in most other countries in Europe [15]. Delays in seeking health services can increase the chance of spreading the virus to others and can result in more severe cases presenting to a healthcare facility – hence the high rates of Covid-19 in these countries.

Excesses in circulatory and respiratory diseases, neoplasms and transport account for most of the non-Covid-19 excess in 2020 but there was a large component left in some countries not accounted for in 2021, in particular in Croatia, Cyprus, Malta and Turkey. In 2021 the non-Covid-19 excess was double the sum of the four in Turkey, with significant excess in certain infectious and parasitic diseases (ICD-10: A00-B99) and in ill-defined and unknown causes of mortality (ICD-10: R96-R99) (p<0.001). For the latter it had the highest rate of any country and just behind Czechia and Serbia in 2020, while in most countries rates were positive but low both years. The Netherlands also had significantly high rates of unknown causes of mortality. Croatia had highly significant positive rates of excess diabetes mortality in both 2020 and 2021 (p<0.001) and while in most countries rates were positive, they were low. Cyprus, Croatia and Czechia had the highest mean endocrine mortality rates in EU pre-Covid-19. Unhealthy diets, tobacco smoking, alcohol consumption, low physical activity in Croatia and a high proportion of obese people have been reported [16]. These factors may have been exacerbated by government stringency measures during the pandemic. Malta is the only country to have significant positive excess mortality due to dementia (ICD-10: F01-F03) both years with rates almost double the next largest country. In other countries, some were positive and some negative. The major causes of disease in Malta are circulatory, respiratory and then Alzheimer’s and other dementias [17]. While rates for Alzheimer’s decreased slightly during the pandemic in Malta this was not so for dementias. A possible explanation is that their very high population density together with government lockdowns led to increased stress levels.

Cyprus in both 2020 and 2021 and Bulgaria and Latvia in 2020 had positive excess neoplasms mortality rates while it was negative or else near zero in all other countries. From Table S1, Covid-19 other rates are not an explanation for excess neoplasm rate in Cyprus. This asks how does Cyprus differ from other countries? It is reported Cyprus, had an initial success in containing the epidemic by implementing strict and early travel restrictions and lockdowns (it had third highest maximum SI in 2020 and seventh highest in 2021). This quick response probably reflects the fact that the public health system was already in a poor state and was not equipped to deal with a large number of cases [18]. In addition, Cyprus had the highest rate of male smokers in the study followed closely by Greece and Latvia. This may have been exacerbated in Cyprus (and to a lesser extent Latvia) during the pandemic, as due to strict lockdown measures smoking rates may have increased. In Belgium, Ireland and France the non-Covid-19 excess mortality is lower than the sum of the four both in 2020 and 2021. The countries with statistically negative non-Covid-19 excess in 2021 are precisely the countries that have statistically significant negative excess respiratory mortality in 2021 indicating fewer deaths due to regular influenza etc. [19].

As expected, excess mortality due to transport was negative in all countries except Poland in 2020 and Estonia, Latvia and Slovenia in 2021 (in the case of Estonia it may point to it having the lowest mean SI in 2020 and 2021) but none were significantly so. This may be due to factors such as accidents that did not occur due, for example, to restrictions on commuting or travel during the lockdown periods. The HDI was positively associated with excess transport mortality suggesting greater road use and car ownership in wealthier countries, although CEEC’s had the highest transport accident rates pre-Covid-19. There is some evidence that the strictest stringency measures were imposed in several eastern European countries during the pandemic thereby reducing transport mortality.

The HDI featured as an explanatory variable in the regression model in all excess deaths due to specific causes (apart from neoplasms), was negatively associated with them and accounted for a large proportion of the variation explained. This is an important health-economic variable that seems to be a good indicator of pandemic preparedness. The proportion of female smokers was positively associated with CID and with excess neoplasms in 2021. This result with CID was also found in [5], who noted the proportion of female smokers is associated with poorer countries which have fewer resources to deal with the pandemic. Median age was negatively associated with excess respiratory and neoplasm mortality and is to be expected as these affect mostly older age groups. Vaccination rates were negatively associated with overall excess, excess respiratory, excess circulatory, CID2021 mortality and a borderline association also with Covid-19 other, most of these to be expected. The proportion age 70 or older was positively associated with excess transport but since excess transport was negative or very low for all countries this association is of little importance.

Stringency variables and vaccination rate variables were all positively correlated in 2021 indicating countries with responsible public health policies had both high vaccination and stringency rates.

Note that mortality for the two main causes of death in Europe was lower than expected in 2020 and 2021 in most countries. Without a specific reason, such as new effective treatments for lung or breast cancer, new drugs for cardiovascular diseases or chronic obstructive pulmonary disease that were effective and widespread at European level at the same time, implemented at the beginning of the pandemic, one might assume that these negative excess deaths were classified as “comorbidities” or underlying causes of death, in death certificates. However, we have no data to justify this.

### Indications for the future

For the four causes of mortality studied excess respiratory diseases stand out as having the greatest number of significantly negative countries particularly in 2021. There are indications the latter is linked to the stringency index and suggests the adoption of face masks and similar measures during a flu season might be worthwhile [20–21]. An unusual wave of childhood pneumonia emerged in China in May 2024, that the WHO attributed to an immunity gap in a cohort of children who were isolated, resulting in large outbreaks once exposure to pathogens returned. Strict pandemic restrictions led to a weakened population immunity—a trend also seen in the UK and USA in 2022 [22]. In Ireland, that has the highest respiratory mortality rates pre-Covid-19 in the EU and had the lowest excess respiratory mortality rates during the pandemic, during the 2022/2023 season, excess pneumonia and influenza mortality was reported over four consecutive weeks [23].

Studies in England and Wales [19,24] have shown a substantial reduction in presentations to hospitals with acute cardiovascular conditions during the pandemic.

It is also reported the Covid-19 pandemic disrupted access to cancer treatment [25–26].

Although we found only weak evidence for the effectiveness of government control policies in this study, social-control measures are still highly relevant in the fight against Covid-19 [27–28].

It is clear from the results on CEEC’s including Croatia, Cyprus, Malta, Turkey [29] and also results in Ireland that the pandemic affected non-Covid-19 mortality rates in different ways that were particular to the country involved. A significant excess in any cause of death, either positive or negative is a cause for concern and a flag for increased rates in the future. Preparing for a future pandemic will require solutions to health problems particular to each country as well as tackling the social determinants of health. For CEEC’s countries the HDI is an important indicator of these problems. The IMF [30] stated, “internationally, strong multilateral cooperation is essential to overcome the effects of the pandemic, including to help financially constrained countries facing twin health and funding shocks, and for channelling aid to countries with weak health care systems”.

Note that in this study using the average numbers of deaths from past years might underestimate the total expected numbers because of population growth or aging but this is unlikely to affect results in any major way as the average is based on just four past years.

## Data Availability

The data that support the findings of this study are openly available in the sources listed in the Methods Section above.

## Acknowledgements

We would like to thank Norman Noah for his advice at the outset of this project.

## Ethical Approval

Not applicable.

## References

1. Bwire G. (2020) Coronavirus: Why Men are More Vulnerable to Covid-19 Than Women? Springer Nature Comprehensive Clinical Medicine; 2:874– 876. doi:10.1007/s42399-020-00341-w

2. Rossen LM, et al. Excess all cause mortality in the USA and Europe during the COVID 19 pandemic, 2020 and 2021. Scientific Reports 2022; 12, 18559. doi:10.1038/s41598-022-21844-7.

3. Eurostat. EU excess mortality above the baseline in May 2023. Published online: 14 July 2023. https://ec.europa.eu/eurostat/web/products-eurostat-news/w/ddn-20230714-2

4. The Organization for Economic Cooperation and Development (OECD)/European Union (EU). COVID 19 mortality and excess mortality. Health at a Glance: Europe 2022: State of Health in the EU Cycle, OECD Publishing, Paris. doi:10.1787/507433b0-en

5. Kelly G, Petti S, Noah N. Covid-19, non-Covid-19 and excess mortality rates not comparable across countries. Epidemiology and Infection 2021;149:e176. doi:10.1017/S0950268821001850.

6. Eurostat. Circulatory diseases, cancer: 54% of all EU deaths in 2021. Published online: 25 March 2024. https://ec.europa.eu/eurostat/web/products-eurostat-news/w/ddn-20240325-2

7. Eurostat database (https://ec.europa.eu/eurostat/data/database). Accessed 15 April 2024.

8. Our World in Data (OWID) database. (https://ourworldindata.org/coronavirus-source-data). Accessed 17 May 2024.

9. Hale T, et al. (2021) A global panel database of pandemic policies (Oxford COVID-19 Government Response Tracker). Nature Human Behaviour; 5: 529–538 doi:10.1038/s41562-021-01079-8

10. The Organization for Economic Cooperation and Development (OECD). Central and Eastern European Countries (CEECs) Definition. *OECD Glossary of Statistical Terms*. OECD Publishing Paris 2008. doi:10.1787/9789264055087-en.

11. Whittaker C, et al. Under-reporting of deaths limits our understanding of true burden of covid-19. British Medical Journal 2021; 375:n2239. doi: 10.1136/bmj.n2239.

12. Beltramo G, et al. (2021) Chronic respiratory diseases are predictors of severe outcome in COVID-19 hospitalised patients: a nationwide study. European Respiratory Journal. Published online: December 2021; 58: 2004474021. doi:10.1183/13993003.04474-2020.

13. Eberhardt N, et al. (2023) SARS-CoV-2 infection triggers pro-atherogenic inflammatory responses in human coronary vessels. Nature Cardiovascular Research; 2: 899–916. doi:10.1038/s44161-023-00336-5.

14. Paul O, et al. (2022) Pulmonary vascular inflammation with fatal coronavirus disease 2019 (COVID-19): possible role for the NLRP3 inflammasome. Respiratory Research; 23: doi:10.1186/s12931-022-01944-8.

15. Džakula A, et al. (2022) A comparison of health system responses to COVID-19 in Bulgaria, Croatia and Romania in 2020. Health Policy; 126: 456–464. doi:10.1016/j.healthpol.2022.02.003.

16. Matassi VG, Rogić A. (2023) Mortality trends in Croatia in the first two decades of the 21st century. Geoadria; 28:97–126. doi:10.15291/geoadria.4094

17. World Health Organization (WHO). Health data overview for the Republic of Malta. Published online: 20 May 2023.https://data.who.int/countries/470

18. Stiftung FE. The impact of the Covid-19 crisis on divided Cyprus. Published online: April 2020. https://library.fes.de/pdf-files/bueros/zypern/16785.pdf.

19. Office of National Statistics. Analysis of death registrations not involving coronavirus (COVID-19), England and Wales: 28 December 2019 to 10 July 2020. Published online: 2 September 2020. https://www.ons.gov.uk/peoplepopulationandcommunity/birthsdeathsandmarriages/deaths/articles/analysisofdeathregistrationsnotinvolvingcoronaviruscovid19englandandwales28december2019to1may2020/28december2019to10july2020

20. Fitz-Simon N, et al. Understanding the role of mask-wearing during COVID-19 on the island of Ireland. Royal Society Open Science 2023;10: 221540. doi10.1098/rsos.221540

21. Zhu D, et al. Social distancing in Latin America during the Covid-19 pandemic: an analysis using the stringency index and Google Community Mobility reports. Journal of Travel Medicine, December 2020;27: taaa125. doi:10.1093/jtm/taaa125.

22. Anon. (2024) Patterns of respiratory infections after COVID-19 [Editorial]. The Lancet Respiratory Medicine; 12:1.

23. Central Statistics Office Ireland. Measuring Mortality Using Public Data Sources 2019-2023 (October 2019 - June 2023). Published online: 04 October 2023. https://www.cso.ie/en/releasesandpublications/fp/fp-mpds/measuringmortalityusingpublicdatasources2019-2023october2019-june2023/

24. Wu J, et al. (2021) Place and causes of acute cardiovascular mortality during the COVID-19 pandemic. Heart; 107:113–119.

25. American Association for Cancer Research. AACR Report on the Impact of COVID-19 on Cancer Research and Patient Care. Published online 9 February 2022: https://www.AACR.org/COVIDReport

26. Islam JY, et al. Characteristics of patients with cancer and COVID-19 who discontinued cancer treatment. Journal of the American Medical Association Network Open 2024;7: e2411859. Published online: 23 May 2024. doi:10.1001/jamanetworkopen.2024.11859

27. Bilinski A, Emanuel EJ. (2020) COVID-19 and excess all-cause mortality in the US and 18 comparison countries. Journal of the American Medical Association; 324:2100–2102. doi:10.1001/jama.2020.20717

28. Lai S, et al. (2020) Effect of non-pharmaceutical interventions to contain COVID-19 in China. Nature; 585: 410–413. doi:10.1038/s41586-020-2293-x.

29. Hajdu T, Krekó J, Tóth CG. Inequalities in regional excess mortality and life expectancy during the COVID 19 pandemic in Europe. Nature Scientific Reports 2024; 14:3835. doi:10.1038/s41598-024-54366-5

30. International Monetary Fund (IMF). World Economic Outlook, April 2020: The Great Lockdown. Published online: April 2020. https://www.imf.org/en/Publications/WEO/Issues/2020/04/14/weo-april-2020.

