## Supplementary Table S1 and text for "Excess respiratory, circulatory, neoplasm and other mortality rates during the Covid-19 pandemic in the EU and their implications"

Supplementary Material

Results.

Covid-19 identified (CID).

From Table S1, Belgium and Slovenia have the highest rates of CID in 2020, while it is the CEEC’s countries (apart from Turkey) that have the highest rates in 2021. CID was positively correlated with excess respiratory 2021 (0.54, p=0.0012), excess circulatory 2021 (0.61, p=0.0002), but not with excess neoplasms or excess transport. In a stepwise regression analysis in 2020, sd SI was positively (p=0.0413) associated with CID (R^2^=0.14), and in 2021, female smokers (p<0.005) and max SI 2021 (p=0.0916) were positively associated while max vaccination rate (p=0.0097) and life expectancy (p<0.0001) were negatively associated (R^2^=0.86). CID in 2021 was negatively correlated with mean vaccination rate (0.69, p<0.0001), sd vaccination rate (0.77, p<0.0001) and max vaccination rate (0.75, p<0.0001). CID was not correlated with stringency statistics in either 2020 or 2021 though all were positive. CID in 2021 was correlated with sd SI 2020 (0.446, p=0.009) and with max SI 2020 (0.38, p=0.027).

Covid-19 non-identified plus other (Covidother)

Belgium had an extremely high rate of Covidother in 2020 in comparison to other countries and it is again comparatively high in 2021 (Table S1). Serbia also had high rates for both years but in general rates are low for all countries in comparison to CID rates. Covidother was not correlated with excess respiratory, excess circulatory, excess neoplasms or excess transport in either 2020 or 2021. In a stepwise regression analysis in 2020, mean SI 2020 was positively (0.31, p=0.0995) associated with Covidother (R^2^=0.09), and in 2021, proportion of hospital beds (p=0.0129) was positively associated (R^2^=0.20). Covidother was negatively correlated in 2021 with mean vaccination rate (0.42, p=0.01), and max vaccination rate (0.46, p=0.0077) and was not correlated with stringency statistics but all were slightly negative. Covidother in 2021 was not correlated with stringency 2020 statistics.

There were no significant correlations between Covid-19 not identified (CNID) and Covid-19 other (CO) and excess mortality rates.

Supplementary Table S1: Covid-19 (identified CID, otherwise Covidoth) standardised mortality rates per 100,000 population in 32 European countries in 2020 and 33 in 2021 †.

| Country | CID2020 | Covidoth2020 | CID2021 | Covidoth2021 |
| --- | --- | --- | --- | --- |
| Austria | 68.46 | 2.93 | 83.51 | 2.04 |
| Belgium | 140.46 | 41.02 | 70.50 | 13.49 |
| Bulgaria | 105.37 | 8.40 | 365.17 | 8.37 |
| Croatia | 107.13 | 0.29 | 203.07 | 0.95 |
| Cyprus | 19.45 | 0.00 | 86.89 | 0.26 |
| Czechia | 108.11 | 1.81 | 242.68 | 10.85 |
| Denmark | 18.9 | 0.40 | 25.48 | 0.43 |
| Estonia | 14.31 | 0.14 | 125.61 | 0.80 |
| Finland | 8.66 | 0.50 | 15.17 | 0.22 |
| France | 74.99 | 11.76 | 76.21 | 1.07 |
| Germany | 38.95 | 1.28 | 71.42 | 1.70 |
| Greece | 38.73 | 0.06 | 127.08 | 0.12 |
| Hungary | 94.35 | 1.85 | 255.6 | 2.59 |
| Iceland | 11.51 | 0.40 | 1.70 | 0.00 |
| Ireland | 53.32 | 1.62 | 84.87 | 0.36 |
| Italy | 94.77 | 5.95 | 81.95 | 0.57 |
| Latvia | 34.57 | 0.19 | 214.71 | 0.65 |
| Liechtenstein | 130.32 | 0.00 | 52.63 | 4.46 |
| Lithuania | 76.07 | 0.14 | 236.79 | 0.55 |
| Luxembourg | 95.1 | 4.45 | 83.21 | 2.77 |
| Malta | 43.57 | 0.00 | 43.89 | 0.00 |
| Netherlands | 108.1 | 16.64 | 116.18 | 1.84 |
| Norway | 8.68 | 0.04 | 16.9 | 0.72 |
| Poland | 116.95 | 4.19 | 255.73 | 11.68 |
| Portugal | 58.33 | 0.94 | 102.58 | 0.23 |
| Romania | 90.80 | 1.23 | 222.52 | 0.22 |
| Serbia | 124.44 | 20.79 | 381.16 | 18.85 |
| Slovakia | 86.98 | 0.00 | 314.74 | 2.16 |
| Slovenia | 161.73 | 0.00 | 133.1 | 7.17 |
| Spain | 112.19 | 25.82 | 73.26 | 1.18 |
| Sweden | 85.85 | 4.05 | 49.18 | 0.47 |
| Switzerland | 104.04 | 1.90 | 66.34 | 0.77 |
| Turkey | N/A | N/A | 156.98 | 0.00 |
| † *, **, *** denotes statistical significance at the 0.05, 0.01 and 0.001 level respectively. | | | | |
